# Implementation of the 2021 molecular ESGO/ESTRO/ESP risk groups in endometrial cancer

**DOI:** 10.1101/2021.03.25.21254244

**Authors:** Sara Imboden, Denis Nastic, Mehran Ghaderi, Filippa Rydberg, Franziska Siegenthaler, Michael D Mueller, Tilman T Rau, Elisabeth Epstein, Joseph W. Carlson

## Abstract

**Introduction:** In 2021, a joint ESGO/ESTRO/ESP committee updated their evidence-based guidelines for endometrial cancer, recommending a new risk grouping incorporating both clinicopathologic and molecular parameters. We applied the new risk grouping and compared the results to those of the prior 2016 clinicopathologic system.

**Materials and methods:** We classified molecularly a cohort of 604 women diagnosed with endometrial cancer using immunohistochemistry for TP53 and MMR proteins on a tissue microarray, as well as Sanger sequencing for *POLE* mutations. These results, combined with clinicopathologic data, allowed the patients to be risk grouped using both the new 2021 molecular/clinicopathologic parameters and the prior 2016 clinicopathologic system. In addition, clinical treatment and outcome data were collected from medical records.

**Results:** The application of the 2021 molecular markers shows Kaplan-Meier curves with a significant difference between the groups for all survival. Molecular classification under the 2021 guidelines revealed a total of 39 patients (39/594, 7%) with a change in risk group in relation to the 2016 classification system: the shift was alone due to either P53abn or *POLEmut* molecular marker. In order to ensure correct 2021 molecular risk classification, not all patients with endometrial cancer need a molecular diagnostic: 386 (65.0%) cases would need to be analyzed by TP53 IHC, only 44 (7.4%) by MMR IHC and 109 (18.4%) *POLE* sequencing reactions.

**Conclusion:** Application of the 2021 molecular risk groups is feasible and shows significant differences in survival. IHC for TP53 and MMR and applying POLE sequencing is only needed in selected cases and leads to shifting risk groups both upward and downward for a sizeable number of patients.

## INTRODUCTION

In January 2021 the European Society of Gynecological Oncology (ESGO), the European Society for Radiotherapy and Oncology (ESTRO), and the European Society of Pathology (ESP) published updated guidelines for risk group determination in endometrial cancer, integrating both molecular diagnostics and clinicopathologic variables, with the goal of improving patient treatment[1]. These molecular prognostic risk groups represent a revolutionary change in the management of women with endometrial cancer (EC) and will require a total reorientation of how these patients are diagnosed and treated. Prior to the issuance of these guidelines, prognostic risk groups were based on postoperative clinicopathologic findings such as tumor histology, stage of disease, grade, and lymphovascular space invasion (LVSI) [2–5]. The new 2021 molecular risk groups are a hybrid of clinicopathologic findings and molecular markers. The new risk grouping serves as the basis for patient management, especially the use and type of adjuvant therapy. However, implementation of the 2021 molecular risk groups carries with it numerous challenges. These include, but are not limited to, (1) introduction of molecular testing for a relatively common tumor that is treated at both specialized cancer centers and smaller, more community-based, centers, (2) interpretation of molecular diagnostic results by clinical teams that may not have encountered them previously, and (3) movement of patients between risk groups in sometimes surprising ways, which may cause worry and concern by the patient and care team.

The molecular understanding of EC has seen an incredible evolution over the past decade. In 2014, The Cancer Genome Atlas (TCGA) [6] developed an integrated genomic classification of endometrial cancer (EC), dividing cases into four genomic subgroups: (1) copy number high “serous-like,” (2) copy number low “endometrioid-like,” (3) polymerase epsilon (*POLE*) ultramutated, and (4) microsatellite unstable “MSI (hypermutated)”, with each group exhibiting a different progression-free survival. Since then, numerous research groups have explored ways to introduce a clinically applicable molecular-based classification system for EC[7–15]. These molecular markers have been evaluated in isolation, as well as in classification schema, in order to compare them with traditional clinicopathologic based systems. Finally, in the 5^th^ addition of the WHO Classification of Female Genital Tumors, published in September 2019, the molecular classification is integrated, finalizing the definite place of molecular markers in the diagnosis of EC [16].

The primary objective of the current study was to evaluate the implementation of the 2021 ESGO/ESTRO/ESP molecular risk groups in a large and unselected EC patient cohort. A second objective was to compare the new risk groups with the 2016 clinicopathologic only, non-molecular risk groups. A third objective was to identify challenges to implementation, with the goal of providing concrete guidance to clinics, large and small, implementing this system [2,17].

## MATERIALS AND METHODS

### Patient cohort and clinical data

The study was approved by the local ethics boards in Stockholm and in Bern (2016/362 and 2018-00479, respectively); ethics approval for the bio banking of tissue was granted from the regional biobank review board in each country. All women provided written informed consent for the use of their bio banked tissue and clinical data for research purposes.

This is a retrospective cohort study; the women were selected without reference to tumor type and therefore can be said to represent a population-based cohort of all EC women during the years 2004-2015. A cohort of 349 women diagnosed with EC from Karolinska University Hospital (2011-2015) and 255 women from Bern University Hospital (2004-2015) was established (KImBer cohort). Clinical and radiological data, medical history, and treatment data as well as pathology-associated parameters were obtained through a review of digital medical records and pathology reports. Follow-up data on recurrence and survival are available through standardized databases and follow-up controls in both clinics. All pathology slides were reviewed by reference pathologists (Karolinska cases, DN, JC, and Bern cases, TR) to confirm grade and histotype.

### Molecular classification: 2021 ESGO/ ESTRO/ESP

Molecular analysis was applied following the WHO Classification of Tumours, 5th Edition, Volume 4: Female Genital Tumours. Cases were classified as *POLE*mut, MMRd, NSMP, and p53abn [7–10,18–21]). We constructed a TMA (tissue microarray) upon which we performed immunohistochemistry for p53 and MMR proteins MLH1, MSH2, MSH6, and PMS2. Mismatch repair protein deficiency (MMRd) was defined as loss of nuclear staining in at least one out of the four MMR proteins. P53 aberrant protein expression (p53abn) was defined as either complete loss of nuclear protein expression or strong homogenous nuclear overexpression.

### *POLE* sequencing

To identify *POLE* (ultramutated) tumors, DNA was isolated from two 1 mm core punches of formalin-fixed paraffin-embedded tumor tissue using a commercial DNA extraction kit (PerkinElmer chemagen Technology, chemangic protocol name: DNA formalin-fixed paraffin-embedded external lysis VD101124.che) according to the manufacturer’s protocol. All EC cases were analyzed for mutations of *POLE* gene (NM.006231) exons 9-14 by Sanger sequencing. Details of primers and interpretation of mutations has been published previously[15]. A tumor was considered *POLE* mutated (*POLE*mut) if sequencing proved the existence of a hotspot mutation in the exonuclease domain *POLE*, as described in our publication on *POLE*mut tumors [15] and verified by Gilks et al [22].

### Multiple classifiers

Analyzing tumors for p53, MMR and *POLE* can reveal defects in multiple molecular markers. These tumors were classified as recommended in the 2021 ESGO/ESTRO/ESP system (i.e., first *POLE*mut, then MMRd, then p53abn).

### Prognostic risk grouping

The patients were divided into two prognostic risk groups: (1) addition of the new 2021 ESGO/ESTRO/ESP molecular classification system and (2) the 2016 clinicopathologic system only.

A flow chart was developed, that allowed an identification of each patient into a risk category by using as few supplementary tasting as possible.

## STATISTICAL ANALYSIS

To address the first objective of our study, prognostic risk groups were examined looking at overall survival (OS), disease-specific survival (DSS), and recurrence-free survival (RFS) using both Kaplan–Meier plots with log-rank significance testing and Cox proportional-hazards regression models.

Associations between molecular classifiers and other variables such as demographic and clinicopathological factors were tested using non-parametric tests. T-test and ANOVA were used for continuous variables; log-rank test, chi-square, and Fisher’s exact test were used for categorical variables.

A statistical significance level of 0.05 was used. IBM SPSS software version 24.0 was applied.

## RESULTS

### General clinicopathological characteristics

Initially, 604 women were included in this study. Molecular classification (one or more molecular markers) was not possible for 10 of them; these women were therefore excluded from the analyses. At initial diagnosis, mean and median ages were 66.1/67 (range 31-93) years, and BMI was 29/27.6 (range 16.4-58.6). Only 53 (8.9%) of the women were premenopausal at the time of initial diagnosis. Most of the women were either para 2 (32.5%) or nulliparous (23.6%) (range 0-9).

### Application of Molecular Markers

Results of the molecular classification and associations with clinicopathological characteristics are summarized in Table 1.

**Table 1.** Association between molecular markers, demographic and clinicopathologic factors.

| Variable |  | Molecular classifier |  |  |  |  |
| --- | --- | --- | --- | --- | --- | --- |
|  | Total<br>N (%) | MMRd | POLEmut | P53adn | NSMP | p-value<br>association |
| **No. Of patients (%)** | 594 (100) | 199 (33.5) | 38 (6.4) | 86 (14.5) | 271 (45.6) |  |
| **Age** | | | | | | $P=0.001$ |
| Mean ±SE | 66±0.4 | 67.2±0.7 | 60.1±1.4 | 67±1 | 65.8±0.6 | |
| Median | 67 | 68 | 60.5 | 67 | 66 |  |
| **BMI mean±SE** | 29.1±0.3 | 28.5±0.5 | 27.3±1.0 | 29.3±0.9 | 29.7±0.5 | $p=0.68$ |
| Missing | 106 | 35 | 2 | 18 | 51 |  |
| **Grade (%)** | | | | | | $p=0.00$ |
| G1 | 226 (38.0) | 59 (29.6) | 13 (34.2) | 14 (16.3) | 140 (51.7) |  |
| G2 | 202 (34.0) | 88 (44.2) | 9 (23.7) | 12 (13.9) | 93 (34.3) |  |
| G3 | 166 (27.9) | 52 (26.1) | 16 (42.1) | 60 (69.8) | 38 (14.0) |  |
| **Histotype** | | | | | | $p=0.00$ |
| Non-endo | 100 (16.8) | 29 (14.6) | 7 (18.4) | 48 (55.8) | 16 (5.9) |  |
| Endometrioid | 494 (83.2) | 170 (85.4) | 31 (81.6) | 38 (44.2) | 255 (94.1) |  |
| **LVSI** | | | | | | $p=0.00$ |
| Yes | 161 (27.1) | 62 (31.2) | 15 (39.5) | 34 (39.5) | 50 (18.5) |  |
| No | 433 (72.9) | 137 (68.8) | 23 (60.5) | 52 (60.5) | 221 (81.5) |  |
| **FIGO Stage** | | | | | | $p=0.00$ |
| I | 443 (74.6) | 147 (73.9) | 33 (86.8) | 49 (57.0) | 214 (79.0) |  |
| II | 55 (9.3) | 21 (10.6) | 2 (5.2) | 9 (10.5) | 23 (8.5) |  |
| III | 69 (11.6) | 27 (13.6) | 3 (7.9) | 14 (16.3) | 25 (9.2) |  |
| IV | 27 (4.5) | 4 (2.0) | 0 (0) | 14 (16.3) | 9 (3.3) |  |
| **Myometrial invasion** | | | | | | $p=0.08$ |
| intramucosal | 53 (8.9) | 16 (8.0) | 1 (2.6) | 4 (4.7) | 32 (11.8) |  |
| <50% | 305 (51.3) | 100 (50.3) | 23 (60.5) | 39 (45.3) | 143 (52.8) |  |
| >50% | 235 (39.6) | 82 (41.2) | 14 (36.8) | 43 (50.0) | 96 (35.4) |  |
| Missing | 1 (0.2) | 1 (0.5) | 0 | 0 | 0 |  |
| **Lymph node status** | | | | | | $p=0.01$ |
| LN removed | 323 | 84 (31.3) | 22 (8.2%) | 51(19.0) | 111(41.1) |  |
| Pos | 63 (10.6) | 20 (10.1) | 2 (5.3) | 17 (19.8) | 24 (8.9) | $p=0.20$ |
| Neg | 236 (39.7) | 76 (38.2) | 21 (55.3) | 41 (47.7) | 98 (36.2) |  |
| **Adjuvant treat** | | | | | | $p=0.00$ |
| No | 342 (57.6) | 111 (55.8) | 22 (57.9) | 28 (32.6) | 181 (66.8) |  |
| Yes | 239 (40.2) | 80 (40.2) | 16 (42.1) | 57 (66.2) | 86 (31.7) |  |
| Missing | 13 (2.2) | 8 (4.0) | 0 (0) | 1 (1.2) | 4 (1.5) |  |
| **Recurrence** | | | | | | $p=0.00$ |
| Yes | 88 (14.8) | 29 (14.6) | 1 (2.6) | 26 (30.2) | 32 (11.8) |  |
| No | 470 (79.1) | 152 (76.3) | 36 (94.7) | 58 (67.4) | 224 (82.7) |  |
| Missing | 36 (6.1) | 18 (9.0) | 1 (2.6) | 2 (2.3) | 15 (5.5) |  |
| **Survival** | | | | | | $p=0.00$ |
| Alive | 486 (82.1) | 162 (81.4) | 37 (97.4) | 60 (69.7) | 227 (83.8) |  |
| DOD | 57 (9.6) | 15 (7.5) | 1 (2.6) | 21 (24.4) | 20 (7.4) |  |
| Other cause | 28 (4.7) | 9 (4.5) | 0 | 3 (3.5) | 16 (5.9) |  |
| Unknown cause | 15 (2.5) | 9 (4.5) | 0 |  | 6 (2.2) |  |
| Treatment | 3 (0.5) | 1 (0.5) | 0 | 2 (2.3) | 0 |  |
| Missing | 3(0.5) | 3 (1.5) | 0 | 0 | 2 (0.7) |  |

Only three clinicopathological factors showed no significant association with the molecular classification (BMI, myometrial invasion and lymph node status: *p* values of 0.68, 0.08, and 0.2 respectively). Or else the groups differ significantly in their clinical features.

### 2021 ESGO/ESTRO/ESP Molecular Prognostic Risk Groups

Application of the 2021 molecular prognostic risk groups to the cohort revealed the distribution in the five risk groups: low risk N=243(40.9%), intermediate risk N=84 (14.1%) high-intermediate N=89 (15.0%) high risk N=148 (24.9%) and advanced N=30 (5.1%). A detailed comparison of the 2021 molecular with the 2016 clinicopathologic only prognostic risk groups is shown as a cross table (Table 2).

**Table 2.** Shifting upward and downward of cases between the 2016 and the 2021 molecular risk groups. The cross table reveals a significant change in the prognostic groups, (p=0,000 Chi-square).

| 2016 Clinicopathologic Risk Groups |  |  |  |  |  |  |  |
| --- | --- | --- | --- | --- | --- | --- | --- |
| 2021 ESGO/ESTRO/ESP<br>Molecular Risk Groups |  | Low risk | Intermed<br>risk | High-<br>intermed<br>risk | High risk | Advanced /<br>metastatic | Total |
|  | Low risk | 221 | 1 | 9 | 12 | 0 | 243 |
|  | Intermed<br>risk | 2 | 82 | 0 | 0 | 0 | 84 |
|  | High-<br>intermed<br>risk | 0 | 0 | 89 | 0 | 0 | 89 |
|  | High risk | 12 | 1 | 2 | 133 | 0 | 148 |
|  | Advanced /<br>metastatic | 0 | 0 | 0 | 0 | 30 | 30 |
|  | Total | 235 | 84 | 100 | 145 | 30 | 594 |

A total of 39 patients changed group in the new classification. In 17 patients, there is a shift upward due to classification as p53abn. In 22 patients, there is a shift downward due to classification as *POLE*mut. The shift of patients between prognostic risk groups, as well as the distribution of the molecular subtypes within the prognostic groups, is shown in Figure 1. Additionally, three patients show advanced, FIGO Stage 3 disease and *POLE* mutation. All three patients had hotspot mutations (S297F, V411L and P286R). In the 2021 molecular risk groups, these patients are not clearly classifiable and were left in the high / advanced risk group.

**Figure 1.**
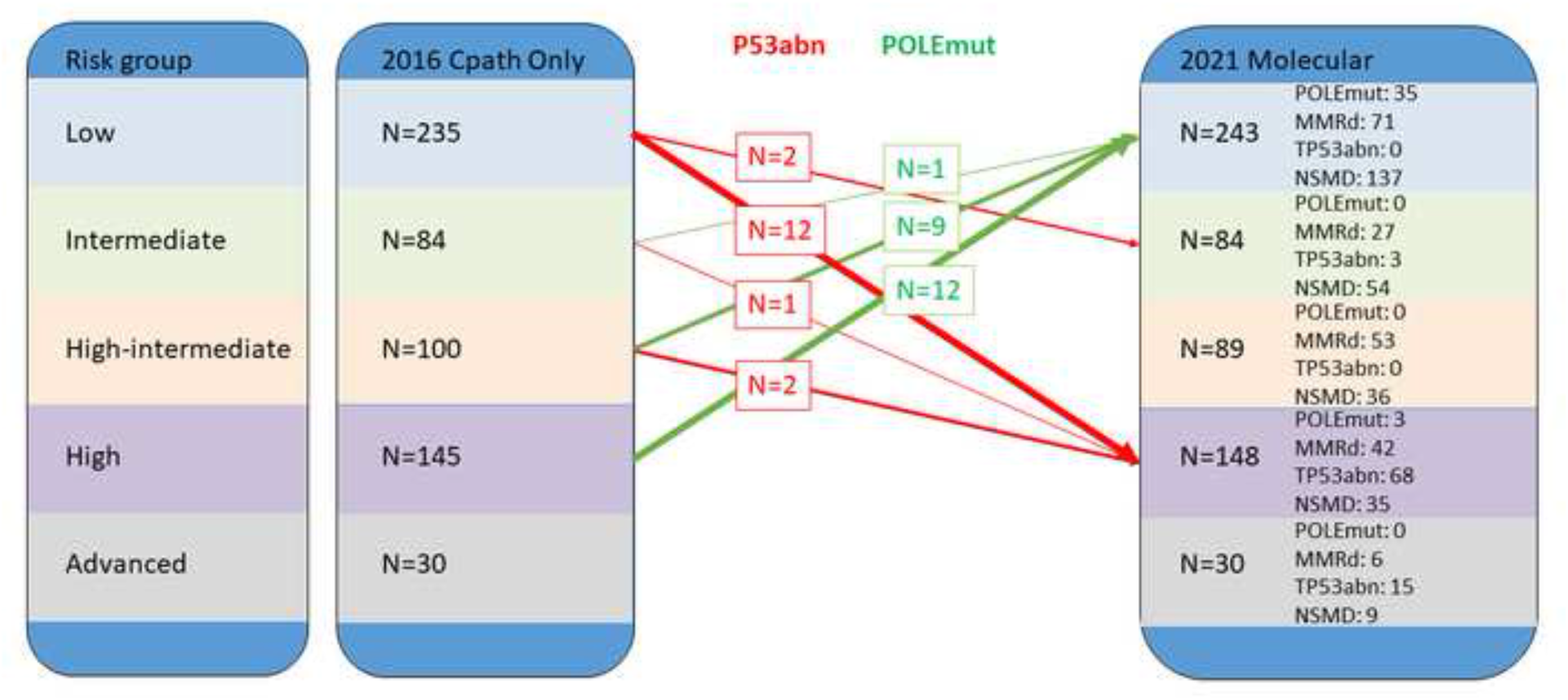
Change between the old and new risk classification. The shift of patients between prognostic risk groups, as well as the distribution of the molecular subtypes within the prognostic groups. Red arrow: p53abn, green: POLEmut

### Double and Triple Classifiers

In this cohort, 47 patients had tumors with multiple molecular aberrations. There were 45 tumors with double aberrations (32 with P53abn and MMRd, 10 with P53abn and POLEmut, 2 with MMRd and POLEmut). There were 2 cases with three aberrations (i.e. P53abn, POLEmut and MMRd).

A total of 126 tumors were P53abn, but of these 10 had a POLEmut and 32 were MMRd (2 both). Thus, only 86 tumors were finally classified as P53abn. Similarly, 204 tumors showed MMRd, but of these 5 had a POLEmut. Thus, only 199 tumors were finally classified as MMRd.

Double and triple classifiers influence the shift of cases between 2021 prognostic risk groups. For example, as described above, a case can be placed in the P53abn classification group only if POLEmut and MMRd have been excluded. In this cohort, 10/594 (1.7%) had both a POLE and p53 mutation, of these 9/10 (90%) were FIGO stage I or II. Similarly, a case can be placed in the MMRd classification group only if a POLEmut has been excluded. In this cohort, 2/594 (0.3%) had both a POLEmut and MMRd.

### Assessment of prognosis

The 2021 molecular risk groups show a significant difference (all log rank *p*= 0.000) among the groups when considering recurrence free survival (RFS), disease specific survival (DSS) and overall survival (OS) (Figure 2).

**Figure 2.**
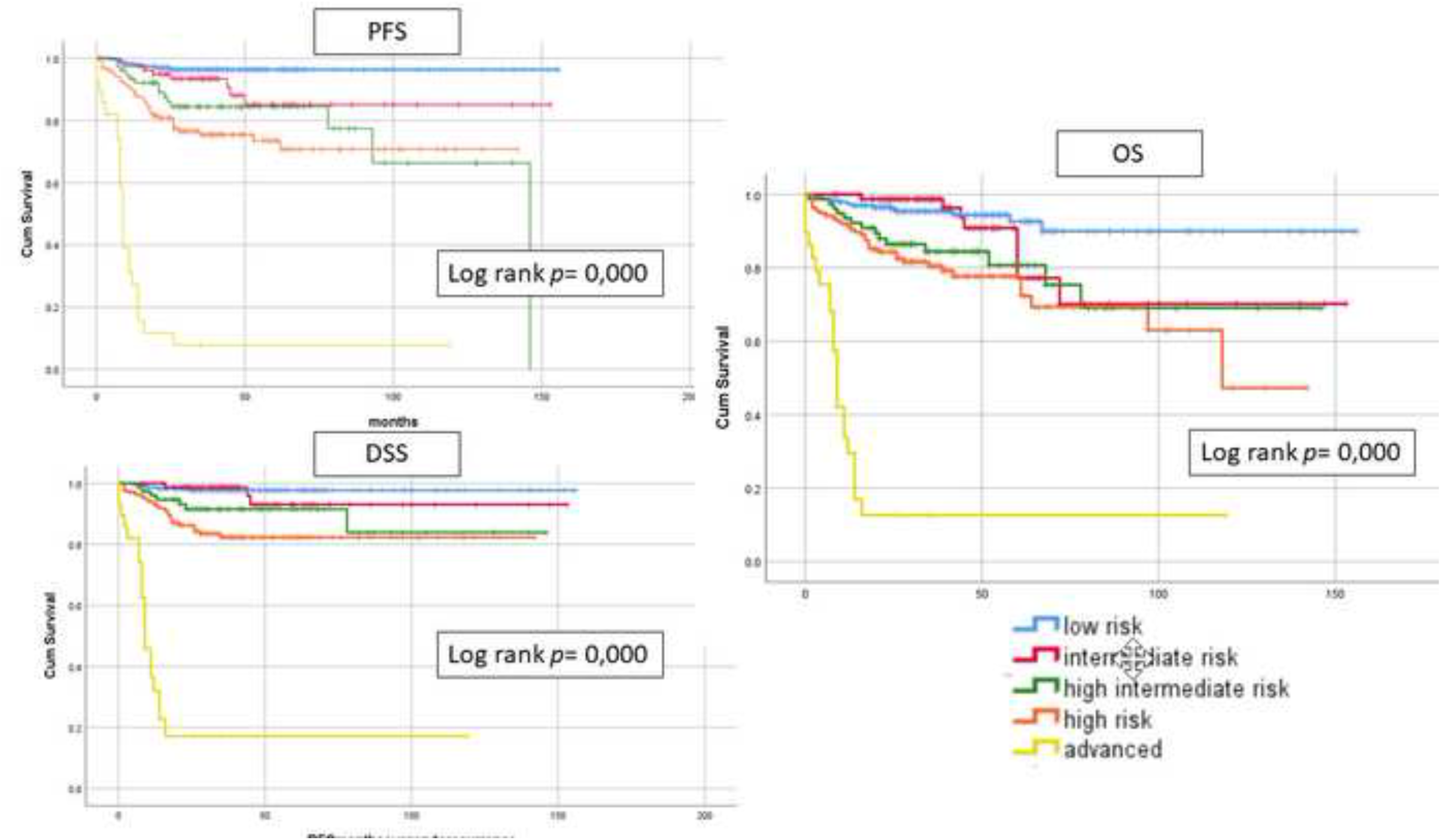
Kaplan-Meier survival curves for PFS for molecular subgrou ps. Kaplan-Meier curves with log-rank test demonstrating the prognosis of patients stratified using the 2021 ESGO/ESTRO/ESP molecular prognostic groups compared to the prior clinicopathologic only risk groups. Comparison performed showing Progression free survival (PFS), Disease specific survival (DSS) and overall survival (OS).

### Implementation of molecular diagnostics

For the 2021 molecular risk grouping, a shift upward is seen only in FIGO Stage I and II tumors classified as P53abn. A shift downward was seen only in tumors classified as *POLE*mut. Therefore, we tested a model where few supplementary diagnostic tests are applied.

In our study we have N=498, 83.3% with all FIGO Stage I and II tumors. In the low risk group with endometrioid Histology, no myometrial invasion, Grade 1or 2 and no LVSI no molecular analysis is needed, since even if they are P53abn the stay in low risk. In the endometrioid cohort therefore, in N=386 (65.0%) patients we need immunohistochemistry for p53. In cases showing an aberrant staining pattern (P53abn) (N=44, 7,4%), immunohistochemistry is needed for MMRd and sequencing for *POLE*mut, to allow correct molecular classification.

In order to identify high risk cases that might potentially be shifted downward due to *POLE*mut classification, an additional 109 (18,4%) cases would need to be *POLE* sequenced.

Therefore, as summarized in Figure 3, in this population based cohort of patients with EC with N=594 patients, in 65% need a staining for P53, 7,4% for MMR Proteins and a total of N=153 (25.8%) need a sequencing for POLEmut (endometrioid and non-endometrioid) to allow a correct 2021 risk grouping.

**Figure 3.**
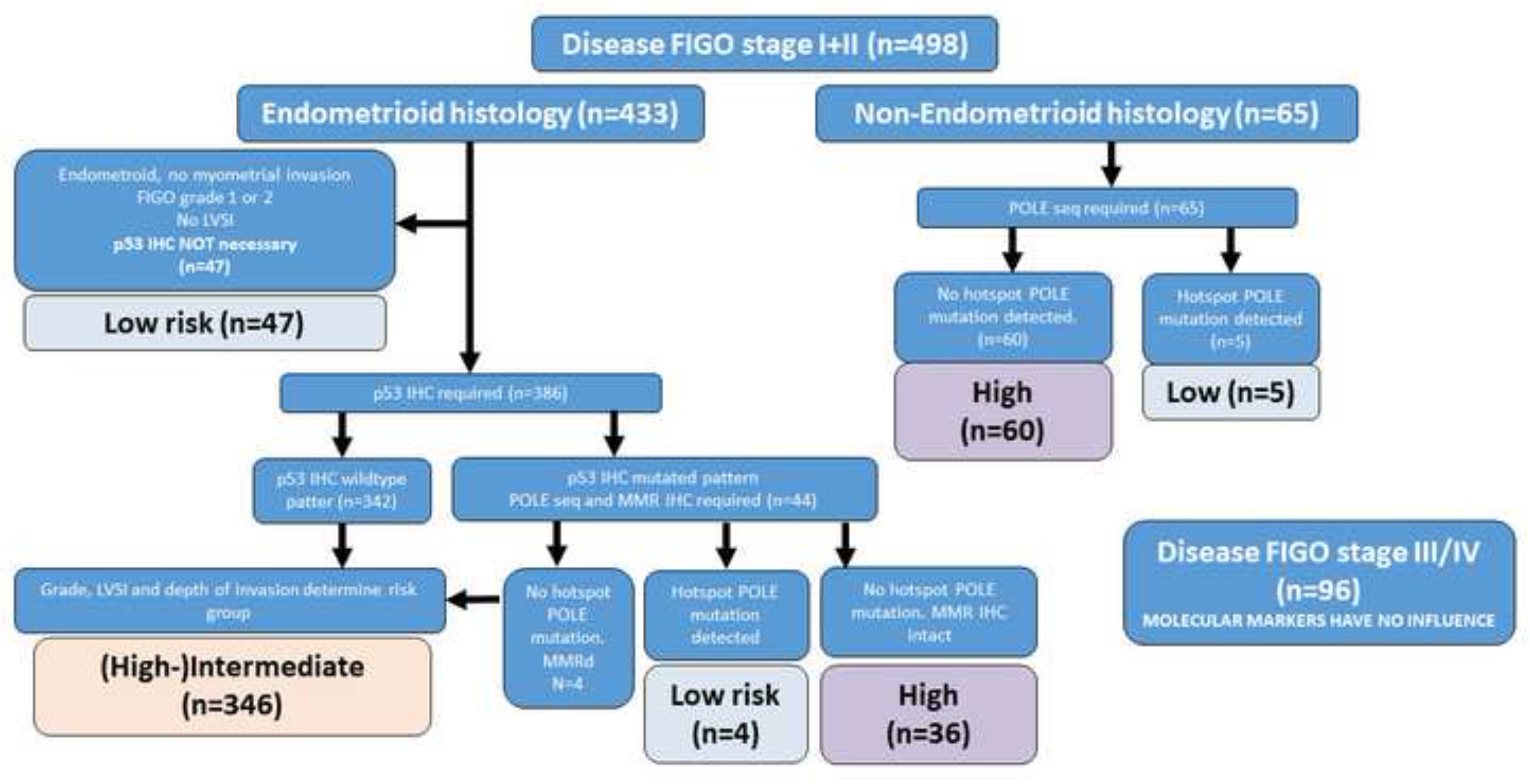
Flow chart with selective analysis for molecular testing. Flow chart, which shows how the patients can be triaged to allow testing for P53, MMR or POLE only when indicated.

## DISCUSSION

In this study, we evaluate the implementation of the 2021 molecular ESGO/ESTRO/ESP prognostic risk groupings in an unselected cohort of endometrial cancer patients. We identify several areas where these groupings differ from their predecessor, and which may cause difficulty upon implementation.

We can confirm that, the implementation of the 2021 molecular prognostic groupings show significant differences in all survival outcomes, which proves the practicability of the new risk grouping. The last adaption with introduction of LVSI was performed 2016 showing similar results in stratifying the risk groups[23,24].

Implementation of the 2021 molecular risk groups on the cohort reveals several important aspects. First, not all P53abn cases are non endometrioid. Indeed, 26 endometrioid FIGO Grade 1 and 2 (low-grade) cases were identified, which would be improperly classified if molecular markers had not been applied. Are all MMRd cases endometrioid? Even here, 29 MMRd cases were judged to be non endometrioid. The results indicate that using histologic subtype to attempt to “triage” cases to molecular subgroups may lead to significant problems with classification and suggest that markers should be applied independent of histology, this was also confirmed in the resent publication by Weigelt et al[25]. Applying TP53 immunohistochemistry on all cases of endometrioid FIGO 1 and 2 would represent a significant change for most pathology labs[26]. In our cohort, fully one-third (26/80, 33%) of P53abn cases would have been missed if endometrioid grade 1 and 2 cases were not stained. Similarly, 15% (29/199) of MMRd cases would have been missed if only endometrioid cases were evaluated for MMRd. Finally, it is important for labs to have access to *POLE* and MMR analyses, because assignment to the TP53 molecular group should occur only if *POLE*mut and MMRd have been excluded. Similarly, assignment to MMRd requires that a *POLE*mut has been excluded. In our algorithm we could show, that not in all cases a molecular diagnostic is needed, rather only in FIGO Stage I and II. By applying IHC for P53 in 65% and POLE sequencing in less than 20% we found a model that is probably more practicable than performing a next generation sequencing for all patients with EC.

Comparing the 2016 clinicopathologic and the 2021 molecular risk classification reveals several interesting aspects. First, 8% of high-risk cases (12/145) downshifted to low-risk after molecular classification due to *POLE*mut. These cases are specifically mentioned in the recommendations as “rare”, but in our cohort they represented the most prevalent reason for risk group shifting[22,27]. In the PORTEC-3 trial, *POLE*mut stage 3 patients had an excellent outcome, but all were treated with external beam radiotherapy. Second, 5% of low-risk cases (12/235) shifted upward to high-risk after molecular classification, due to P53abn. Given that low-risk disease is the most common, this means that potentially 1 out of 20 low-risk cases may be misclassified, and subsequently undertreated, without molecular classification. Again, PORTEC-3 indicates that combined therapy leads to a statistically significant survival advantage for P53abn stage 1-3 [28]. Finally, note that the 2021 recommendations make a difference between P53abn with and without myometrial invasion[1].

Should molecular evaluation be performed preoperatively? Studies have shown that molecular markers performed upon biopsy material show similar results to those performed on hysterectomy material[29]. Indeed, biopsy material can have the advantage of more rapid fixation, potentially allowing for more reliable analysis. However, it is important to note that the 2021 molecular risk groups cannot be determined using biopsy material alone, due to the inclusion of clinicopathologic variables such as FIGO Stage and lymphovascular space invasion. Thus, it is not clear that information beyond histologic tumor type and grade is necessary.

Is it necessary to identify MMRd patients if it does not contribute to their risk grouping? There are several motivations for identifying MMRd patients beyond risk classification, including the desire to identify patients with Lynch syndrome. This is a worthy and important goal for reasons outlined in numerous publications[30,31]. However, from the standpoint of risk classification, MMRd is necessary to correctly classify patients as P53abn. It is important to keep the motivation for the testing clear, so that clinical genetics is consulted and involved in these changes in practice.

Evaluating the minimum testing regime necessary to correctly classify patients revealed that correct classification could be achieved in this cohort with 386/594 (65%) TP53 immunohistochemistry, 44/594 (7%) MMR immunohistochemistry, and 153 (26%) *POLE*mut analyses. Note that additional MMRd analyses may be useful for adjuvant therapy if immunotherapy is potentially indicated.

## CONCLUSION

The 2021 ESGO/ESTRO/ESP molecular risk groups represent a fundamental change in the clinical diagnosis and treatment of endometrial cancer. Implementing and evaluating these changes is a complex task that is only just beginning. Compared to the 2016 clinicopathologic risk groups, 3.7% of high-risk patients were shifted downward to low-risk due to a *POLE* mutation, and 2.9% of low-risk patients were shifted upward to high risk due to a P53abn classification. It is possible to significantly reduce the number of analyses required to implement the classification if resources are limited. Finally, it is important to note that the P53abn group requires exclusion of *POLE*mut and MMRd, making it distinct from prior descriptions of TP53 mutated tumors (where these exclusions were not made). Although no cases were shifted upward or downward due to MMRd, this analysis is useful in identifying women with Lynch syndrome or as a marker for immunotherapy.

## Data Availability

Due to ethical constraints, the individual level cohort data cannot be made publicly available.

## Funding information and Acknowledgments

The funders had no role in study design, data collection and analysis, decision to publish, or preparation of the manuscript. Funding sources are: Bernese Cancer League (https://bern.krebsliga.ch/), Swiss National Science Foundation (IZSE70_177073)http://www.snf.ch/; Avtal om Läkarutbildning och Forskning (ALF), ALF-Stockholm County (grant no 550411)https://ki.se, Cancer research funding from ‘Radiumhemmet’ Stockholm, Sweden (grant no 154112) https://www.rahfo.se, Magnus Bergvalls Stiftelse, Cancerfonden, and Foundation for clinical-experimental cancer research.

## Authors’ contributions

SI, EE, DN, TTR and JWC designed the study protocol and data collection, FR, FS and SI collected, cleaned and analysed the data, SI, DO, DN and JWC performed statistical analysis, MG performed the molecular analysis and analysed the mutations, all authors drafted and revised the paper.

## Abbreviations

EC: endometrial cancer
LVSI: lymphovascular space invasion
POLE: polymerase epsilon
TGCA: The Cancer Genome Atlas
TMA: tissue microarray
OS: overall survival
DSS: disease-specific survival
RFS: recurrence-free survival
IHC: Immunohistochemistry
ESGO: European Society of Gynecological Oncology
ESTRO: European Society for Radiotherapy and Oncology
ESP: European Society of Pathology

## Highlights

- Application of the 2021 ESGO/ESTRO/ESP molecular risk groups is feasible and shows significant differences in survival.
- Immunochemistry for TP53 and MMR and applying POLE sequencing is only needed in selected cases.
- The shift in the risk groups are done by P53and and POLEmut classifications in a sizeable number of patients.

